# The role of personality traits in COVID-19 vaccination: A socio-cultural and gender-stratified analysis in Japan

**DOI:** 10.64898/2026.10.04.26363389

**Authors:** Masamitsu Takamatsu, Mikiko Tokiya, Megumi Hara

## Abstract

**Background:** COVID-19 vaccination uptake is influenced by various sociodemographic and psychological factors. Previous studies have suggested that the relationship between personality traits and vaccination is context-dependent. However, this association has not been investigated in Japan—a unique sociocultural context characterized by historically low vaccine confidence but exceptionally high COVID-19 vaccine uptake. We examined the association between the Big Five personality traits and completion of the primary COVID-19 vaccination series, exploring potential gender–personality interactions.

**Methods:** A web-based, cross-sectional survey was conducted among 7,210 Japanese adults (aged 20–85 years) in December 2021. Personality was assessed using the Japanese version of the Ten-Item Personality Inventory. Multivariable Poisson generalized estimating equations were used to calculate adjusted prevalence ratios (PRs) and assess gender–personality interactions.

**Results:** Higher extraversion and lower openness were significantly associated with vaccination completion across genders. A significant gender–agreeableness interaction was observed: agreeableness (PR = 1.016, 95% CI: 1.008–1.024) was positively associated with vaccination completion among men but not women. A similar, though only marginally significant, interaction pattern was observed for neuroticism (PR = 1.011, 95% CI: 1.004–1.019 among men).

**Conclusions:** Personality traits act as modest but meaningful, gender-specific psychological modifiers of vaccination behavior in Japan. Public health campaigns should transition from traditional “one-size-fits-all” messages to personality-tailored and gender-responsive communication strategies to optimize vaccine uptake and bolster psychological resilience in future pandemics.

## 1. Introduction

One year after the declaration of coronavirus disease 2019 (COVID-19) as a pandemic by the World Health Organization (WHO), multiple vaccines against COVID-19 were approved and deployed worldwide, starting with the United Kingdom on December 8, 2020 [1, 2]. Japan began its COVID-19 vaccine rollout on February 17, 2021, and rapidly increased the coverage despite its delayed approval and introduction [3–5]. By December 31, 2021, 80.4% of Japan’s total population had completed the primary vaccination series. Notably, this rate was significantly higher than the global coverage of 48.6% and the highest among the Group of Seven (G7) nations, whose vaccine uptake rates varied around 70% [6].

COVID-19 vaccination uptake is influenced by many factors at the individual and social levels [7, 8]. Many studies, including those conducted in Japan, have explored associations between personality traits and health behaviors, such as adherence to precautionary measures against COVID-19[9–13]. As COVID-19 vaccines were being developed and rolled out, numerous studies, particularly those focusing on Western countries, investigated the associations between vaccination and personality traits utilizing the five-factor model of personality, commonly known as the Big Five personality traits [14–16]. A recent systematic review by Bleidorn et al. [17] found that high levels in agreeableness, extraversion, and openness and low level in neuroticism are associated with positive attitudes toward vaccination. Additionally, the review found that extraversion is associated with vaccination more strongly in non-Western samples than in Western ones, whereas agreeableness exhibited a stronger association in Western samples than in non-Western ones [17]. These findings suggest that the relationship between personality and vaccination is context-dependent and varies based on the sociocultural background.

Vaccine hesitancy is a serious public health threat that was listed as a critical agenda on the WHO’s list of Ten Threats to Global Health in 2019 [18]. In a 2020 global study, Japan was found to have low vaccine confidence [19]. In fact, Japan has experienced a human papillomavirus (HPV) vaccine scare, which prevented active government recommendations in national programs until recently [20]. Thus, COVID-19 vaccination in Japan serves as a useful case study with a unique paradox. Despite previously reported low vaccine confidence, the Japanese public rapidly embraced COVID-19 vaccines once they became available, leading to a high national coverage rate. This paradox underscores the need to examine the psychological mechanisms underlying vaccination decisions. In the health belief model (HBM), personality falls under sociopsychological variables, which act as modifiers indirectly influencing health-related behaviors [21]. However, to date, no study has examined the association between personality traits and the COVID-19 vaccination uptake in Japan. Furthermore, although previous studies have reported gender difference in COVID-19 vaccination intention in Japan [22], the interaction between gender and personality in the context of COVID-19 vaccination remains poorly understood.

Therefore, this study examined the association between the Big Five personality traits and completion of the primary COVID-19 vaccination series in Japan approximately one year after vaccine rollout. Given Japan’s sociocultural emphasis on social harmony and conformity, we hypothesized that higher agreeableness would be positively associated with vaccination completion. In addition, considering the gender-stratified social pressures in Japan, we explored whether these associations differed by gender.

## 2. Methods

### 2.1 Study design, participants, and ethical approval

A web-based, cross-sectional questionnaire survey was conducted between December 20 and 22, 2021. We used a Japanese online research provider (Macromill, Inc., Tokyo, Japan) to assess COVID-19 vaccination status and the Big Five personality traits. Participants were recruited from a voluntary panel of approximately 1.3 million registered members. Reward points exchangeable for commercial products or services were provided upon survey completion.

Invitations were emailed to men and women aged 20–85 years. Based on previous studies, the required sample size was calculated as 6800, assuming a significance level of 0.05, power of 0.8, vaccination coverage of 80%, and an adjusted odds ratio of 1.13 [23–25]. Although we aimed to recruit 7000 participants, 7210 individuals were ultimately enrolled.

The survey was administered in Japanese. The invitation email clearly stated the study purpose and indicated that the anonymized data may be published. Participation was voluntary, and web-based informed consent was obtained from all participants. Data were hosted on a secure platform, and all responses were mandatory for submission. This study was conducted in accordance with the Declaration of Helsinki and approved by the Ethics Committee of Saga University (approval number: R2–24).

### 2.2 Measurement methods

#### 2.2.1 Sociodemographic and clinical factors

The following sociodemographic and clinical data were collected: gender, age group (20–29, 30–39, 40–49, 50–59, 60–69, or 70–85 years), marital status (married or not married), annual household income (< JPY 4 million or ≥ JPY 4 million), educational attainment (high school graduate or higher education), and the presence of any underlying health conditions.

#### 2.2.2 COVID-19 vaccination status

Participants reported the number of COVID-19 vaccine doses received: none, one dose, two doses, or three doses. In Japan, the administration of the third dose (first booster) began on December 1, 2021, for priority groups, such as healthcare workers and older people (≥ 65 years) who had completed their primary series at least six months prior. As all vaccines approved in Japan at the time required a two-dose primary series, for this analysis, participants who had received two or more doses were defined as having completed their primary vaccination series.

#### 2.2.3 Big Five personality traits

The Big Five personality traits were assessed using the ten-item personality inventory (TIPI) [26]. Despite its brevity, the TIPI is recognized as a reliable and comprehensive instrument based on the following key principles: (1) broad coverage of the Big Five personality traits, (2) inclusion of factors representing both poles of each dimension, (3) exclusion of evaluatively extreme items, (4) avoidance of simple negation items, and (5) minimization of redundancy in items [26]. This study employed the validated Japanese version of the inventory (TIPI-J) [27].

The TIPI-J consists of five item pairs representing five dimensions: (1) extraversion, reflecting extraverted and enthusiastic, in contrast to reserved and quiet (reverse-scored); (2) agreeableness, reflecting sympathetic and warm, in contrast to critical and quarrelsome (reverse-scored); (3) conscientiousness, reflecting dependable and self-disciplined, in contrast to disorganized and careless (reverse-scored); (4) neuroticism, reflecting anxious and easily upset, in contrast to calm and emotionally stable (reverse-scored); and (5) openness, reflecting open to new experiences and complex, in contrast to conventional and uncreative (reverse-scored).

Responses were rated on a seven-point Likert scale (1 = strongly disagree; 7 = strongly agree). For each dimension, the score of the second (negative-pole) item was reverse-coded, and the scores of the two items were summed. This resulted in a score range of 2 to 14 for each personality dimension.

### 2.3 Statistical analysis

#### 2.3.1 Preliminary analysis

A cross-sectional analysis was conducted to examine the association between personality traits and COVID-19 vaccination status. The primary outcome was the completion of the primary vaccination series, defined as having received two or more vaccine doses. The preliminary analysis proceeded in two steps. First, participant characteristics were compared among three vaccination status groups (0 doses, 1 dose, and ≥ 2 doses) using a chi-squared (χ^2^) test for categorical variables. Second, the TIPI-J scores were compared between the completed (≥2 doses) and not completed (0 or 1 dose) groups using the Wilcoxon rank-sum test.

#### 2.3.2 Multivariable analysis

To directly estimate prevalence ratios (PRs) for a common binary outcome and avoid the overestimation of associations that is inherent in odds ratios, we employed Poisson generalized estimating equations (GEE) with a log link function. This method was used to evaluate the associations between personality traits and vaccination completion while adjusting for covariates. Analyses were performed using three hierarchical models. A crude model was used to examine the unadjusted associations between personality traits and vaccination completion. Model 1 was adjusted for sociodemographic and clinical covariates, namely, gender, age, marital status, annual household income, educational attainment, and underlying health conditions. Model 2 was built upon Model 1 by incorporating significant interaction terms. We evaluated potential interactions between personality traits and sociodemographic and clinical factors. Significant interactions identified in this step (e.g., gender × agreeableness) were subsequently included in Model 2 to assess effect modification. All estimates were presented as PRs with corresponding 95% confidence intervals (CIs). Participants with missing information on annual household income (n = 1,615) were excluded from the multivariable analyses (Models 1 and 2, the gender-stratified analyses, and the sensitivity analyses), leaving 5,595 participants for these analyses.

#### 2.3.3 Model fit and distribution selection

To ensure the validity of our statistical approach, we compared the Poisson distribution with a negative binomial distribution. Model selection was based on quasi-likelihood under the independence model criterion (QIC) and its simplified version (QICu) values. In addition, overdispersion was assessed using the ratio between Pearson chi-square and degrees of freedom (χ²/df), with a ratio substantially greater than 1.0 indicating overdispersion.

#### 2.3.4 Sensitivity analysis

As a sensitivity analysis, a multivariable logistic regression was performed to confirm the robustness of our findings. All statistical analyses were conducted using SAS software (Version 9.4; SAS Institute, Cary, NC, USA), with a two-sided significance level set at p < 0.05.

## 3. Results

### 3.1 Participant characteristics according to the number of COVID-19 vaccine doses received

The participant characteristics are presented in Table 1. Unvaccinated individuals were significantly more likely to be young, unmarried, have low annual household income, have low educational attainment, and lack underlying health conditions. From a behavioral medicine perspective, these characteristics represent modifying variables within the HBM that can influence an individual’s internal appraisal of vaccine-related risks and benefits.

**Table 1.**
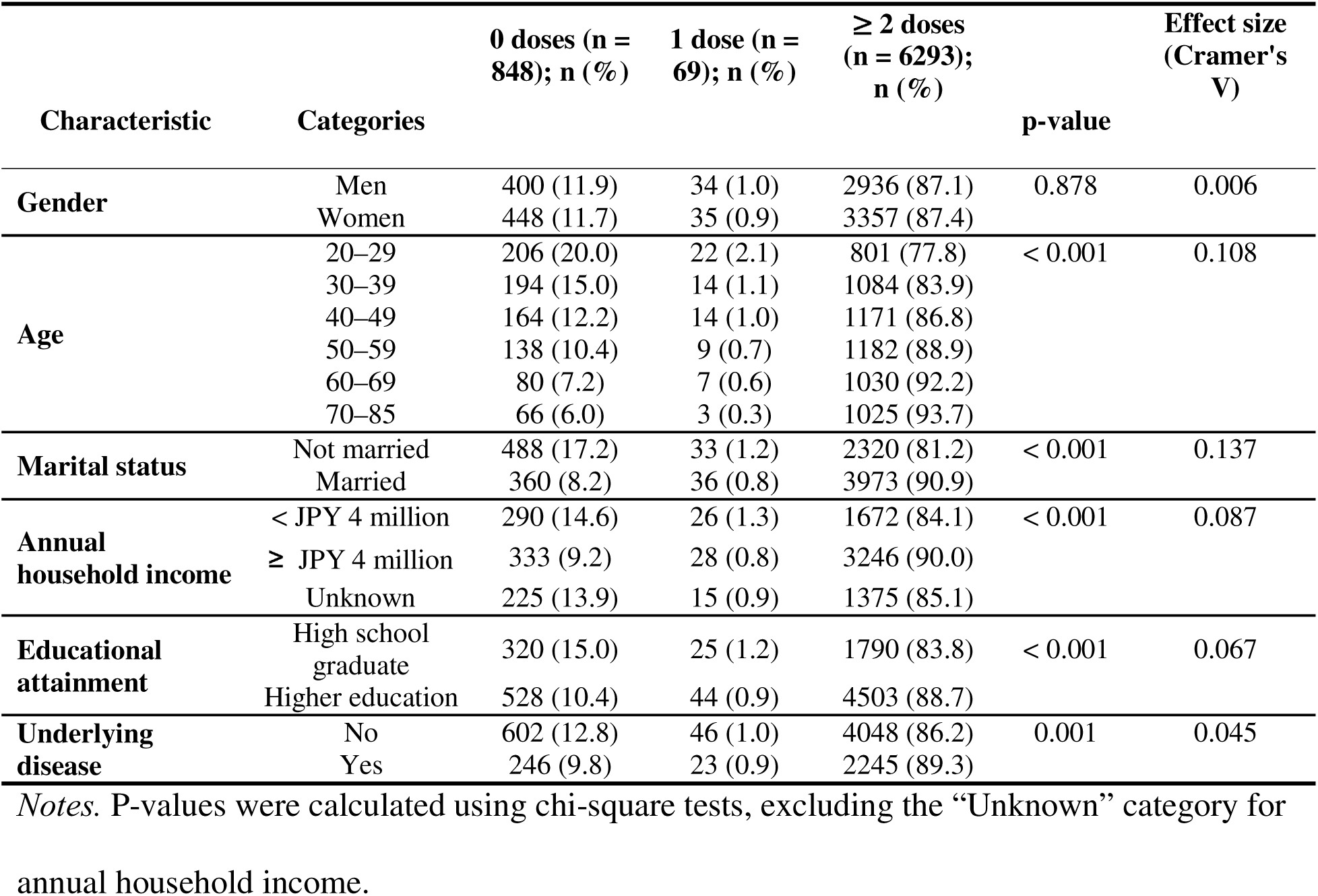
Participant characteristics based on the number of COVID-19 vaccine doses received.

### 3.2 Comparison of the Big Five personality traits according to COVID-19 vaccination completion status

Figure 1 shows the TIPI-J scores based on COVID-19 vaccination completion status. Significant differences were observed in the median scores of agreeableness, extraversion, openness, and conscientiousness. Neuroticism did not show a significant difference between participants who completed the primary series and those who did not. The effect sizes (r) for the Wilcoxon rank-sum tests were as follows: r = 0.038 for extraversion, r = 0.048 for agreeableness, r = 0.025 for conscientiousness, r = 0.013 for neuroticism, and r = 0.038 for openness. Within the HBM framework, these significant differences, particularly higher scores in extraversion and agreeableness and lower scores in openness among vaccinated participants than among non-vaccinated ones, illustrated that personality traits served as foundational psychological modifiers of health behavior.

**Figure 1.**
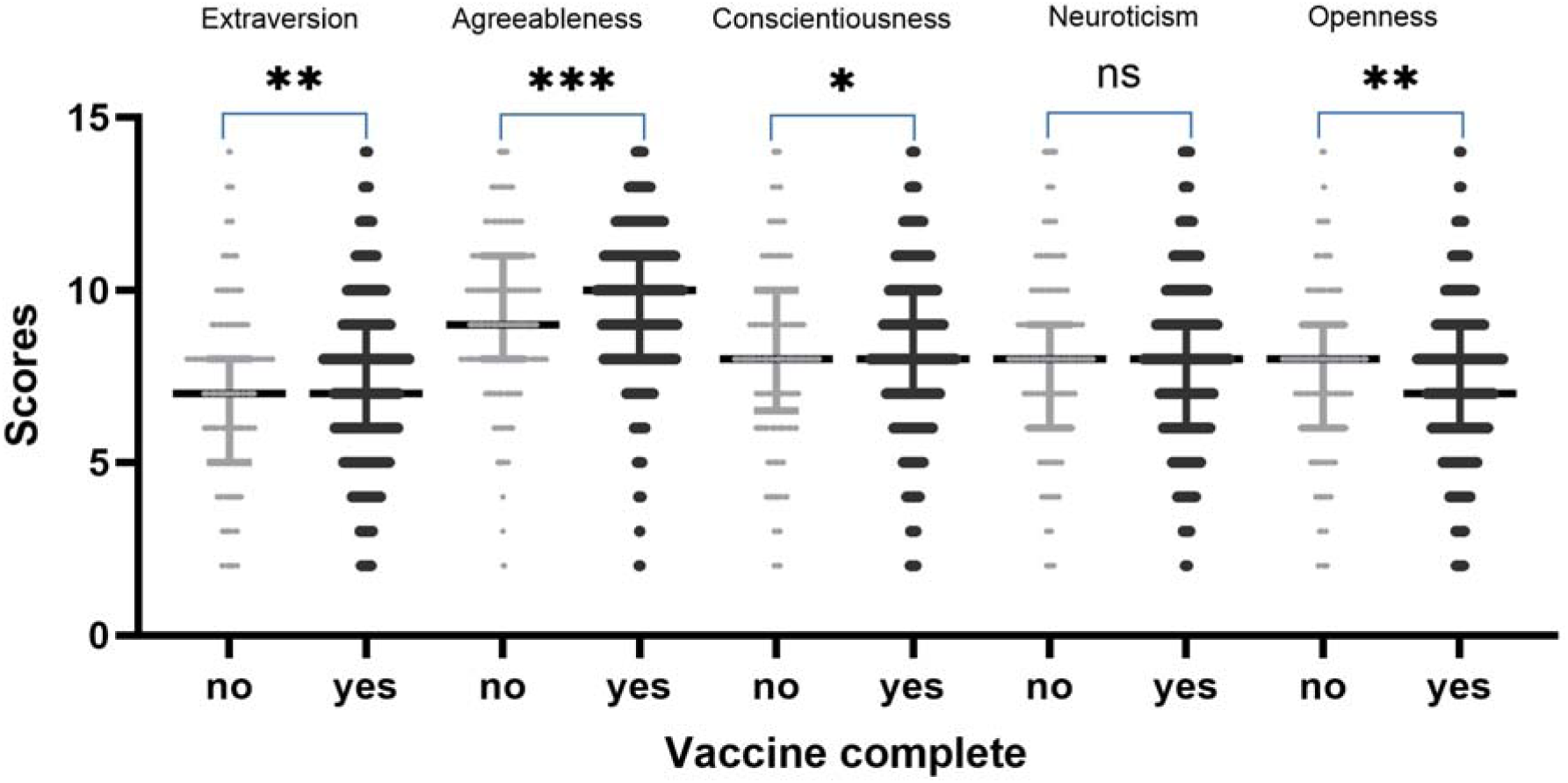
Big Five personality trait scores according to COVID-19 vaccination completion status (n = 7,210). *p < 0.05, **p < 0.01, ***p < 0.001, ns (not significant). Notes. Personality traits were assessed using the TIPI-J. Central horizontal lines represent the median, and error bars denote the interquartile range. Individual data points are displayed as dots. P-values were calculated using the Wilcoxon rank-sum test to compare trait scores between individuals who completed the primary vaccination series (“yes”) and those who did not (“no”).

### 3.3 Model selection and statistical validity

To evaluate factors associated with vaccine completion while adjusting for covariates, we compared Poisson GEE and negative binomial GEE models. The Poisson GEE model was selected for the final analysis because its QIC values were comparable to those of the negative binomial model and it showed stable convergence (Table S1). The Pearson χ^2^/df ratios (Men: 0.124; Women: 0.119) were well below 1.0, indicating no evidence of overdispersion.

### 3.4 Interaction effects and model refinement

The multivariable analysis revealed that higher levels of extraversion, agreeableness, and neuroticism were associated with a higher likelihood of vaccination completion, whereas openness was negatively associated with it. Conscientiousness showed no significant association. Furthermore, a significant interaction was observed between gender and agreeableness, with a marginal interaction between gender and neuroticism (Table 2). From a behavioral perspective, these results represented the extent to which each personality trait functioned as a modifier of the likelihood of completing the primary vaccination series. Based on these findings, gender-stratified analyses were conducted.

**Table 2.**
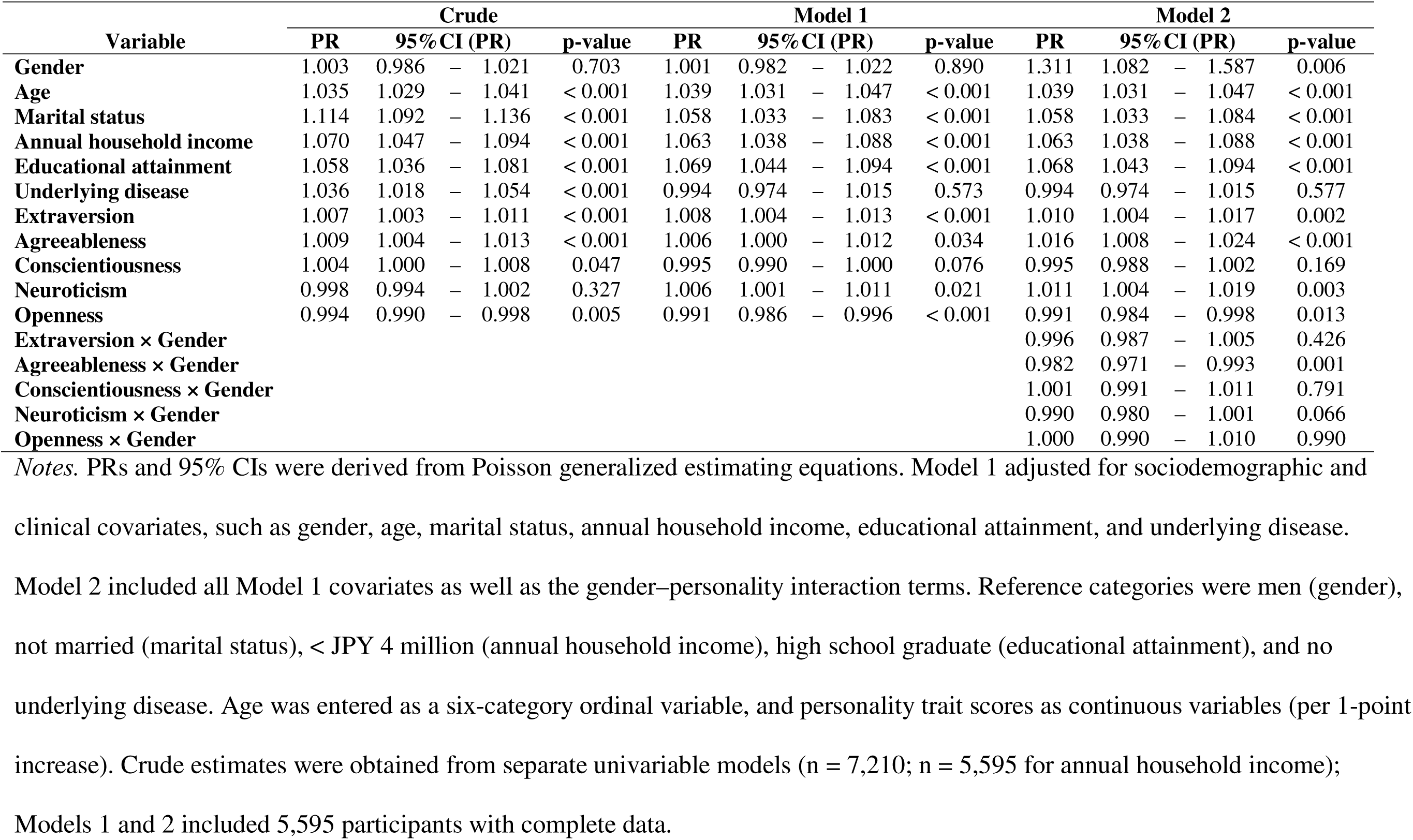
Associations between the Big Five personality traits and vaccine completion.

### 3.5 Gender-stratified analysis of personality traits

The results of the gender-stratified Poisson GEE analysis are presented in Figure 2. Among men, agreeableness and neuroticism were positively associated with vaccination completion, whereas these effects were non-significant among women. Among both men and women, older age, higher education, and higher extraversion were associated with a higher PR for vaccine completion, whereas openness was associated with a lower PR. Interpreted through the HBM, this indicated that gender acts as a major moderator of personality-driven vaccination behavior. It should be noted that the formal interaction term reached statistical significance for agreeableness (interaction p = 0.001) but was only marginal for neuroticism (interaction p = 0.065–0.066), so the neuroticism finding should be interpreted with more caution.

**Figure 2.**
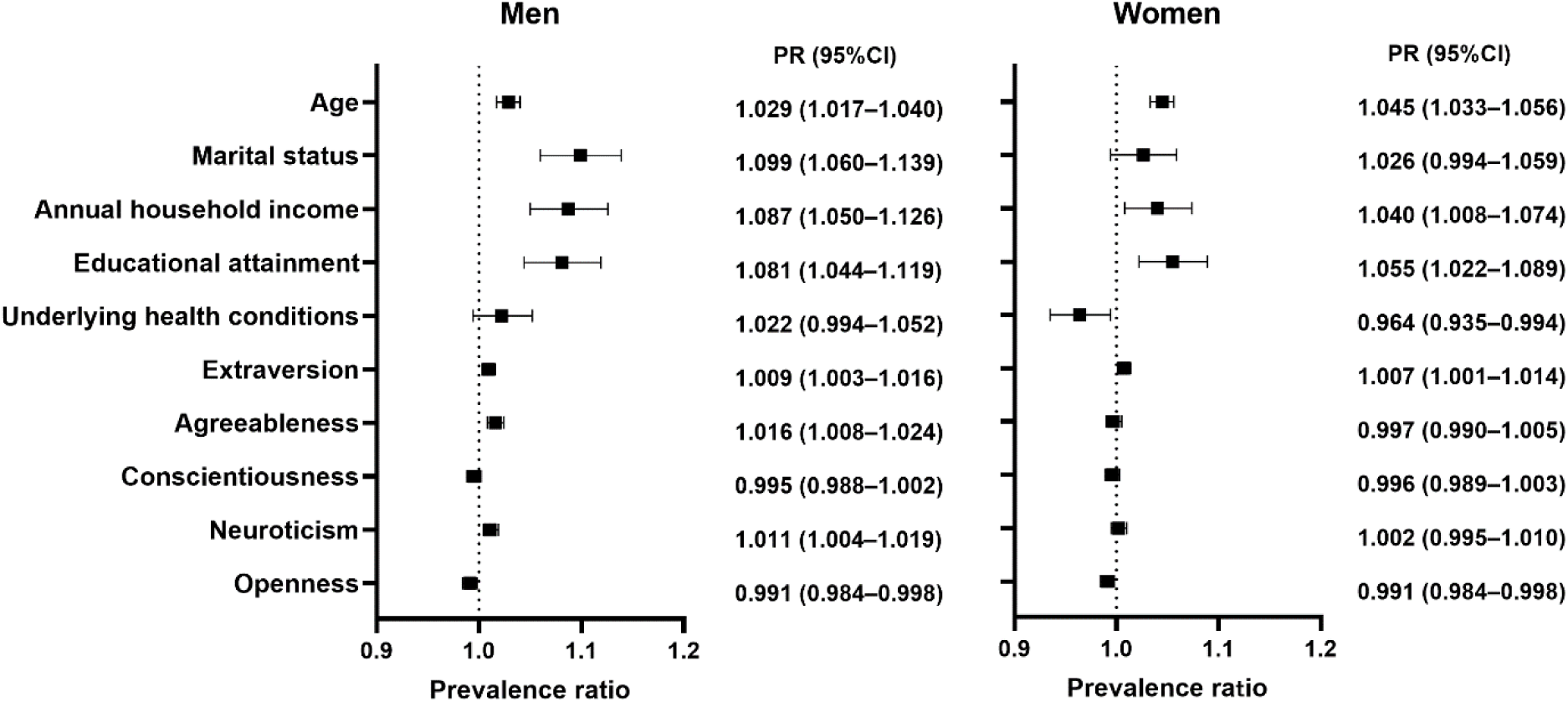
Forest plot of adjusted PRs for gender-stratified primary COVID-19 vaccination completion (n = 5,595; men, n = 2,842; women, n = 2,753). *Notes*. PRs and 95% CIs were estimated using gender-stratified Poisson generalized estimating equation models with a log link function. Models were mutually adjusted for all Big Five personality traits and further adjusted for age, marital status, annual household income, educational attainment, and underlying health conditions. Black squares and horizontal bars represent adjusted PRs and 95% CIs, respectively. Values to the right of the vertical dashed line (PR > 1.0) indicate a positive association with vaccination completion.

Significant positive PRs for agreeableness, and a statistically weaker pattern for neuroticism, exclusively among men suggest that these traits may function as psychological modifiers that enhance the perceived benefits of social conformity or the perceived threat of the disease considering Japan’s gender-specific social roles. Conversely, the lower PRs for openness among both men and women highlight a unique socio-cultural modifier in Japan, where lower openness, which is potentially linked to higher adherence to established social norms, facilitates vaccination completion.

### 3.6. Sensitivity Analysis

The multivariable logistic regression results were highly consistent with the primary Poisson GEE model. The gender-stratified logistic model confirmed that agreeableness was positively associated with the outcome among men, whereas no such association was observed among women (Table 3). Furthermore, the interaction between gender and agreeableness remained significant (Table 4).

**Table 3.**
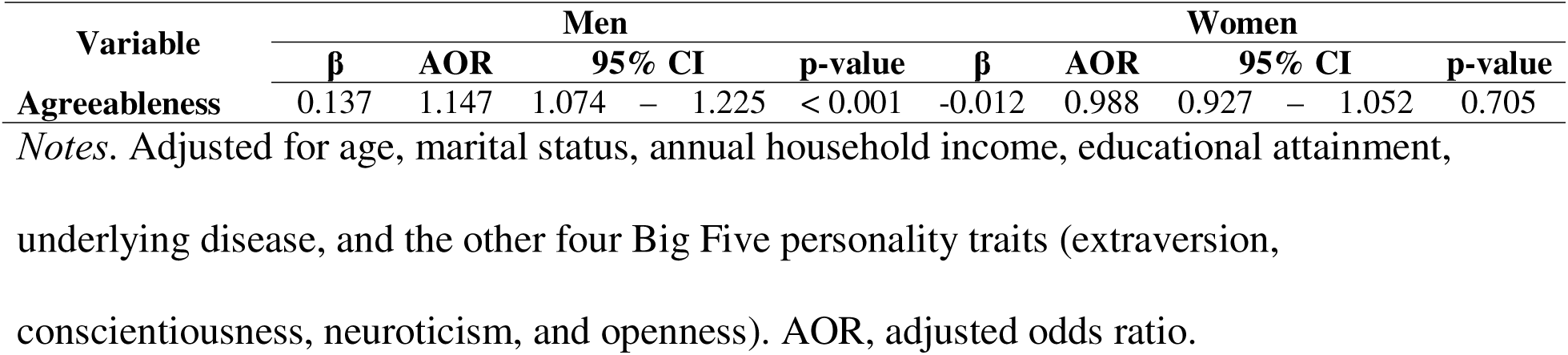
Sensitivity analysis: gender-stratified logistic regression analysis of the association between agreeableness and vaccine completion.

**Table 4.**
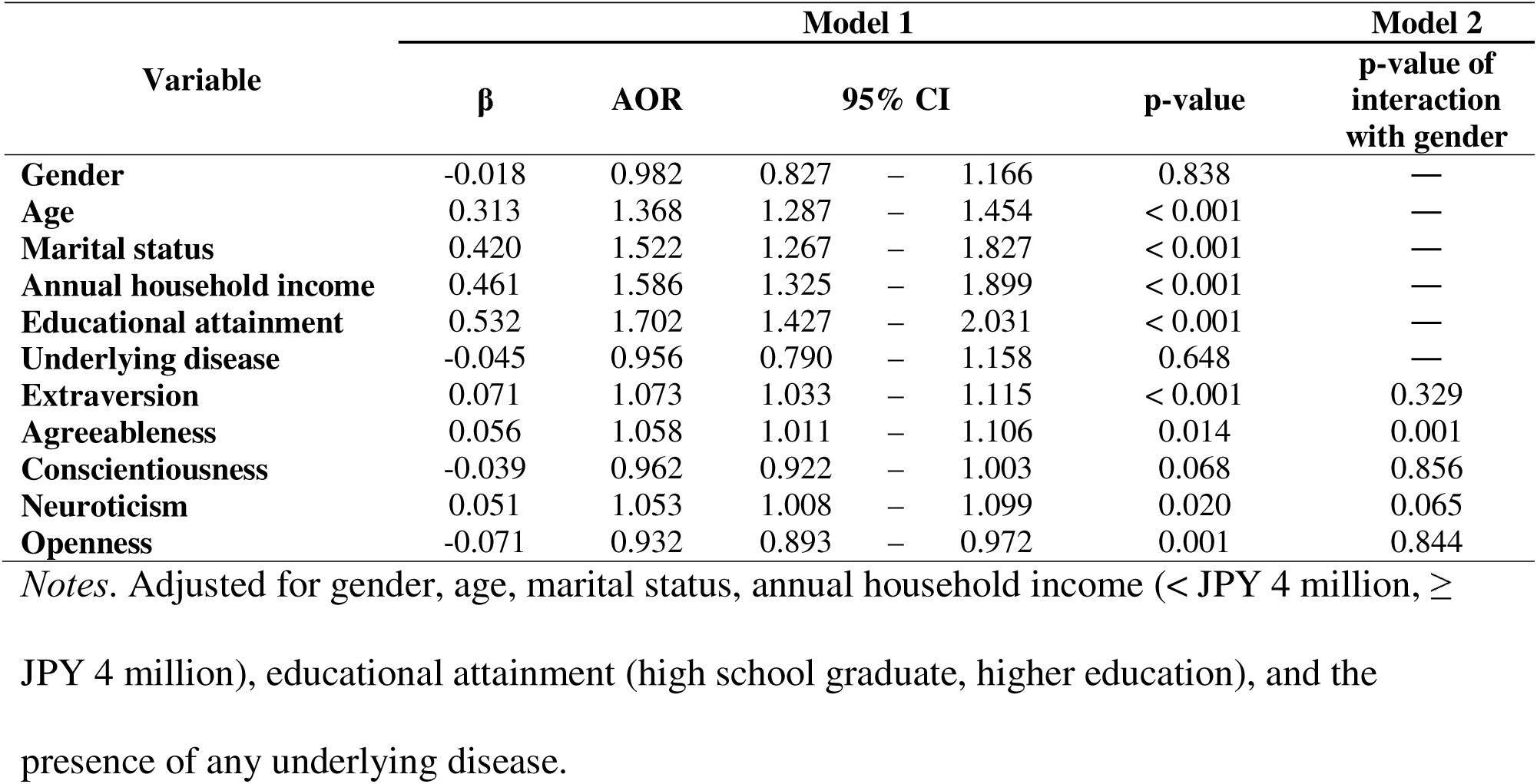
Adjusted odds ratios and interaction effects for vaccine completion estimated using multivariable logistic regression.

The analysis was stratified by gender to examine the moderating effect of gender-responsive social roles on personality traits. Within the HBM framework, these results suggest that for Japanese men, traits such as agreeableness act as psychological modifiers that significantly enhance the perceived benefits or threats associated with vaccination. Conversely, the lack of significant associations among women may reflect a complex interplay between safety concerns (perceived barriers) and health beliefs.

Adjusted odds ratios were estimated using a multivariable logistic regression analysis to assess the robustness of the associations found in the primary Poisson model. The consistency of the significant gender–personality interactions across different statistical frameworks reinforced the interpretation that personality traits serve as stable psychological modifiers of vaccination behavior in the Japanese sociocultural context.

## 4. Discussion

This study investigated the association between the Big Five personality traits and the completion of the primary COVID-19 vaccination series in Japan. Three personality traits, namely, extraversion, agreeableness, and neuroticism, were positively associated with vaccination status. This supports our hypothesis that individuals with higher agreeableness are more likely to be vaccinated than those with lower agreeableness. Our findings provide unique insights, as we assessed vaccination status approximately one year after the initial vaccine rollout and concurrent with the beginning of the booster dose campaign in Japan. The primary series vaccination coverage among the study participants (87.3% for individuals aged 20–85 years) was comparable to the national coverage of approximately 80% at the time.

Our findings regarding agreeableness and extraversion align with the results of a global review by Bleidorn et al. [17], whereas neuroticism and openness exerted effects in the opposite direction from their findings. From the perspective of the HBM, our results suggest that personality traits serve as modifying factors that influence health behaviors and may affect Japanese individuals’ internal perceptions of risks and benefits of COVID-19 vaccination [21]. The positive association with extraversion may be consistent with the concept that individuals who value social engagement are more motivated to complete vaccination, perceiving it as a necessary benefit to resume social activities during a period when social activities are strongly constrained. Unexpectedly, higher neuroticism was associated with greater vaccination completion. This suggests that neuroticism, as a modifying factor, may increase the perceived threat of COVID-19 among anxious individuals, causing the perceived benefits of protection to outweigh the perceived risks, such as side-effect concerns [9, 16]. Although similar findings have been reported in several studies from Europe [28, 29], this requires further investigation. Nevertheless, this is a valuable insight for Japan, which is a society characterized as risk-averse, where public anxiety surrounding the HPV vaccine side effects previously fueled vaccine scares and undermined confidence [20]. In Japan, where vaccination decisions may be influenced by strong perceived social norms [25], individuals with high openness may be less likely to follow the prevailing consensus than those with low openness. This interpretation regarding the openness paradox is speculative, as we did not directly measure perceived norms or information exposure. However, this is consistent with a psychological study suggesting that individuals with high openness think independently from sociocultural norms [30].

In our study, the influence of agreeableness on vaccination status was moderated by gender, whereas neuroticism had marginal interaction with gender. After stratifying the analysis, both traits showed a positive association with vaccination completion only among men, although this pattern should be considered well established for agreeableness and only exploratory for neuroticism, given its marginal interaction. These findings were consistent across statistical frameworks and reflected gender-related differences in vaccine decision-making. A previous systematic review revealed stronger effect of agreeableness among men than among women [17]. Agreeableness among men could have been more directly linked to a decision to vaccinate, perhaps driven by social norms [25]. From the beginning of the pandemic, the male sex was a prominent risk factor for COVID-19 severity [31]. This could have instigated their neuroticism toward vaccination. Several studies have indicated that women tended to report lower COVID-19 vaccine acceptance and higher concern for vaccine safety than men [32, 33], despite perceiving COVID-19 as a serious disease [34]. In the HBM framework, such mixed attitudes, where a high perceived threat is countered by high perceived barriers (safety concerns), could have attenuated or canceled out the influence of personality traits on their final decision [21]. Our findings indicate that personality as a HBM modifying factor is highly contingent upon the sociocultural and gender-dependent context of the individual. This affirms the need for further research on gender-responsive interventions, as highlighted in the WHO immunization agenda 2030 [35].

Although statistically significant, the effect sizes for personality traits were modest relative to several sociodemographic correlates reported in other studies [14, 17, 28]. Sociodemographic factors such as older age, being married, higher household income, and higher educational attainment were positively associated with vaccination completion, which was in line with the findings of previous studies [36, 37]. Although personality traits play a role, public health interventions should continue to prioritize targeting sociodemographic groups with lower uptake, such as younger, single individuals with lower income and education levels. In practice, these insights can help physicians and health workers tailor their communication with potential vaccinees. Specifically, they can emphasize family protection among highly agreeable men and deliver highly reassuring safety data to address the side-effect concerns among those with higher neuroticism. As more evidence is generated in this field, future immunization guidelines may benefit from including practical tips that incorporate personality and gender factors.

The main strength of this study is that it is the first study in Japan to assess the association between COVID-19 vaccination status and the Big Five personality traits, measured using the validated TIPI-J. In addition, our large sample size allowed us to perform robust gender-stratified analyses and reveal gender-specific patterns. However, this study had several limitations. First, potential misclassification could have occurred because vaccination status was self-reported and personality traits were assessed using the abbreviated TIPI-J scale, which has lower reliability than full-length inventories. Second, we did not account for detailed community characteristics, such as urban-rural distinctions, local disease epidemiology, and the dominant political affiliation. In this study, the information on residential areas was collected as the subnational regions, which are groups of prefectures or second administrative levels. These data did not include community characteristics. Third, the use of an online survey panel may have introduced a selection bias. Nevertheless, our sample’s age distribution was consistent with national demographics, and this data collection method is common in the field. Finally, due to the cross-sectional study design, we could not infer causality between personality traits and vaccination behavior. To address these limitations, future research should employ longitudinal designs to establish causal relationships. Additionally, utilizing full-length personality inventories may reveal more nuanced facet-level dynamics than the brief TIPI-J. Future studies may also benefit from utilizing immunization records to minimize self-report bias. Furthermore, they may incorporate multi-level analyses to examine how individual personality interacts with macro-level cultural and environmental factors, such as local disease transmission, in health decision-making.

## 5. Conclusion

This study revealed that personality traits acted as modest but meaningful psychological modifiers of vaccination behavior among adults in Japan. Furthermore, a significant gender difference was observed for agreeableness, with a similar but only marginally significant pattern for neuroticism, alongside broader cultural specificity. These findings suggest that future public health interventions should prioritize personality-tailored and gender-responsive communication over traditional one-size-fits-all strategies. Utilizing the Big Five framework to design stratified vaccine communication may optimize uptake and bolster psychological resilience in future pandemic responses.

## Data Availability

The datasets used and/or analyzed during the current study are available from the corresponding author

## List of abbreviations

AOR: Adjusted odds ratio
CI: Confidence interval
COVID-19: Coronavirus disease 2019
G7: Group of Seven
GEE: Generalized estimating equations
HBM: Health belief model
HPV: Human papillomavirus
JPY: Japanese yen
PR: Prevalence ratio
QIC: Quasi-likelihood under the independence model criterion
QICu: Simplified (corrected) version of QIC
TIPI: Ten-Item Personality Inventory
TIPI-J: Japanese version of the Ten-Item Personality Inventory
WHO: World Health Organization

## Declarations

## Ethics approval and consent to participate

This study was conducted in accordance with the Declaration of Helsinki and approved by the Ethics Committee of Saga University (approval number: R2–24). Web-based informed consent was obtained from all participants.

## Consent for publication

Not applicable.

## Availability of data and materials

The datasets generated and/or analyzed during the current study are not publicly available to protect the privacy of the participants but can be availed from the corresponding author on reasonable request.

## Competing interests

The authors declare that they have no competing interests.

## Funding

This work was supported by a research grant for Research on Emerging and Re-emerging Infectious Diseases, Health and Labor Science Research Grants from the Ministry of Health, Labour and Welfare, Japan (grant numbers: R2-SHINKOGYOSEI-SHITEI-003 and 20HA2001).

## Authors’ contributions

(MH = Megumi Hara; MsT = Masamitsu Takamatsu; MkT = Mikiko Tokiya.) MH: Conceptualization, Methodology, Investigation, Resources, Project administration, Funding acquisition, Writing – review & editing. MkT: Methodology, Formal analysis, Data curation, Writing – original draft, Writing – review & editing, Visualization. MsT: Writing – original draft, Writing – review & editing, Visualization. All authors read and approved the final manuscript.

## Acknowledgements

We would like to thank Editage (www.editage.jp) for English language editing. We would also like to express our sincere gratitude to all the participants who cooperated in the survey and to Miyuki Fuchigami for her administrative support.

## Additional files

**Additional file 1**: Table S1. Comparison of model goodness-of-fit statistics by gender. (DOCX) *This table evaluates the goodness-of-fit of models (Poisson vs. negative binomial distributions) using criteria such as QIC and Pearson* χ*²/df to justify the selection of the final GEE Poisson regression model*.

## Notes

### Competing Interest Statement

The authors have declared no competing interest.

### Author Declarations

Ethics Committee of Saga University gave ethical approval for this work (approval number: R2-24). Web-based informed consent was obtained from all participants.

