## Supplementary Table S1 for "The role of personality traits in COVID-19 vaccination: A socio-cultural and gender-stratified analysis in Japan"

**Table S1: Comparison of Model Goodness-of-Fit Statistics (by Gender)**

| **Sex** | **Distribution** | **QIC** | **QICu** | **Pearson χ2/df** | **Decision** |
| --- | --- | --- | --- | --- | --- |
| **Men** | **Poisson** | **45368.08** | **45367.77** | **0.124** | **Selected** |
|  | Negative Binomial | 45368.11 | 45367.80 | — | Questionable convergence |
| **Women** | **Poisson** | **45915.10** | **45914.01** | **0.119** | **Selected** |
|  | Negative Binomial | 45915.14 | 45914.04 | — |  |

Notes: * GEE: Generalized Estimating Equations. QIC (Quasi-likelihood under the independence model criterion) and QICu (simplified QIC) were used to evaluate and compare the goodness-of-fit for GEE models with different distributions; lower values indicate a better model fit. Pearson chi^2^/df: The ratio of the Pearson chi-square statistic to the degrees of freedom was used to assess overdispersion. Model Selection: The Poisson distribution was selected for the final analysis as it demonstrated stable convergence and comparable fit to the Negative Binomial distribution. Specifically, for the male cohort, the Negative Binomial model showed questionable convergence, leading to the selection of the Poisson distribution to ensure the robustness of the results across both sexes.
